# Linkage and association of preserved cognitive function in the Midwestern Amish at higher genetic risk of Alzheimer disease

**DOI:** 10.1101/2025.01.06.25320073

**Authors:** Daniel A. Dorfsman, Michael B. Prough, Alex Gulyayev, Laura J. Caywood, Jason E. Clouse, Sharlene D. Herington, Susan H. Slifer, Larry D. Adams, Renee A. Laux, Yeunjoo E. Song, Audrey Lynn, Sarada L. Fuzzell, Sherri D. Hochstetler, Kristy Miskimen, Leighanne R. Main, Ping Wang, Yining Liu, Noel Moore, Paula Ogrocki, Alan J. Lerner, Jeffery M. Vance, Michael L. Cuccaro, Jonathan L. Haines, Margaret A. Pericak-Vance, William K. Scott

## Abstract

**Background:** Genetic studies of Alzheimer’s disease (AD) have made remarkable progress in identifying genetic factors that increase the risk of AD. However, the genetic determinants that contribute to preserved cognition at older ages, particularly those that mitigate AD risk, are less understood. In the Midwestern Amish community, certain older adults maintain normal cognitive abilities despite carrying a relatively high genetic risk of AD. Our goal was to identify shared genetic factors among ‘high-risk’ Amish sibships enriched with cognitively unimpaired individuals. We hypothesized that ranking sibships by their mean genetic risk of AD would provide greater evidence of genetic linkage in cognitively preserved sibships at higher risk, identifying genomic regions containing protective loci.

**Methods:** Cognitive status in adult Amish participants (n = 1,855) was evaluated using the Modified Mini Mental Status Exam (3MS). Those aged ≥ 75 with education-adjusted 3MS ≥ 87 were classified as cognitively unimpaired (CU), while those with education-adjusted 3MS < 87 were classified as cognitively impaired (CI). Genome-wide non-parametric linkage analysis was performed on 143 sibships with ≥ 2 CU members. To account for known genetic risk, ordered-subsets analysis (OSA) was used to rank families by the mean AD genetic risk score (GRS) of their CU members using effect sizes for 25 established European AD risk loci. Regions exhibiting significant or suggestive increases in the LOD* score during OSA were further evaluated using single-variant and interaction association tests in a sample of 715 CU and 460 CI participants.

**Results:** Significant evidence of linkage following OSA adjustment was detected on chromosome 2 at SNP rs6719884 (∼ 59.0 Mb) within the lincRNA *LINC01122* (LOD* = 3.08), suggesting a shared genetic factor influencing preserved cognition in high-risk individuals. Additionally, a significant interaction effect was observed on chromosome 12 at SNP rs11063479 (∼ 5.1 Mb) near the *KCNA5* gene. The variant showed increased odds of being cognitively unimpaired in the high-GRS stratum (OR = 2.07) and decreased odds in the low-GRS stratum (OR = 0.70), indicating a potential synergistic effect with genetic risk.

**Conclusions:** Our findings highlight specific genomic regions—particularly on chromosomes 2 and 12—that may harbor genetic loci contributing to preserved cognitive function in aged individuals at high genetic risk for AD. The identification of *LINC01122* and *KCNA5* as candidate genes underscores the potential role of brain-expressed lincRNAs and potassium channel genes in preserving cognitive function. These results contribute to the understanding of cognitive preservation and may inform future research on protective genetic mechanisms against cognitive decline.

## Introduction

Alzheimer’s disease (AD) is the most common cause of dementia in older adults, accounting for at least two-thirds of all cases. ^1^ AD is etiologically complex—its risk of occurrence depends on both genetic and environmental exposures. Twin studies have estimated the heritability of AD to approximately 70%, which indicates a substantial genetic component.^2^ AD is conventionally categorized based on age at symptom onset into early-onset (<65 years) and late-onset (≥65 years).^3^ Between 1% and 5% of AD cases are early-onset (EOAD). While a small proportion of EOAD cases are caused by highly penetrant autosomal dominant variants in *APP*, *PSEN1*, or *PSEN2*., the majority of EOAD cases are sporadic or familial without a clear inheritance pattern. Late-onset AD (LOAD) is more common (>95%), and does not have a Mendelian subset, with risk driven by a combination of genetic and environmental factors. The strongest genetic factor for developing LOAD is *APOE* genotype, which encodes three main isoforms: ε3 (reference), ε4 (risk-increasing), and ε2 (risk-decreasing).^4,5^ Nevertheless, ongoing genome-wide association studies (GWAS) and meta-analyses have steadily added to the growing catalogue of known AD risk variants, most having low-to-moderate effect size.^6,7^ The Midwestern US Amish population studied here is descended from the same Western European populations that comprise the majority of the study samples that identified these genes.

Despite the complex genetic architecture of LOAD, much of the research focus has been on identifying genetic variation associated with an increased risk of AD. However, it is equally worthwhile to prioritize the discovery of genetic variation that associates with preserved cognitive function at older ages, as this might further explain the variability in LOAD risk and illuminate potential new avenues for therapeutic intervention. Recent studies of human aging have found that longevity clusters within families, and that longevous individuals have a greater tendency to avoid age-related disease.^8^ In addition, the Long Life Family Study reported that siblings and offspring of individuals demonstrating ‘exceptional survival’ carried a cognitive advantage when compared to spousal controls.^9^ The familial concordance of superior aging traits such as longevity and cognitive preservation suggest that genetic factors partly drive these traits. Several genetic variants linked to preserved cognitive function, primarily in European-descended individuals, have been reported. The protective effect of the APOE ε2 allele is well established. Compared to the common ε3/ε3 genotype, ε2 reduces the risk of Alzheimer’s disease and significantly lowers the risk relative to ε3/ε4 and ε4/ε4 genotypes.^5^ Additionally, *APOE* ε4 carriers with a specific allele of the Klotho gene, Klotho-VS, have lower odds of developing AD than ε4 carriers without Klotho- VS.^10^ A rare *APP* substitution, p.A673T, initially discovered in the Icelandic population, is protective against Alzheimer’s disease and is also associated with superior cognitive performance in cognitively unimpaired individuals.^11^ Several replicated GWAS variants in genes such as *SORL1, CLU, and EPHA1* also are associated with a reduced risk of AD.^6,7^ When family data are available, integrating linkage and association-based analytical methods may increase the power to detect trait-associated loci. Association studies of complex, late-onset traits can have lower statistical power when looking for multiple alleles slightly more common in unaffected controls. Studies of pedigrees enriched for such alleles may improve statistical power. Based on prior evidence of familial aggregation of both cognitive preservation and AD, we designed a family-based linkage and association study to identify protective variants for AD in the Midwestern US Amish population. The study utilized non-parametric linkage analysis (NPL) to detect significant departures from expected identical-by-descent (IBD) sharing between aged, cognitively unimpaired sibling pairs (sib-pairs) across multiple nuclear families, potentially identifying regions harboring protective variants. These regions were subsequently examined for association using case/control analysis, while adjusting for familial relationships. This approach may be particularly powerful in population isolates, such as the Amish, due to their closed structure, extensive genealogical records, and adherence to a traditional lifestyle, which results in a homogenous population with low environmental variability.^12^ Previously, our group has conducted linkage and association studies in Amish communities from Ohio and Indiana, identifying regions associated with AD and successful aging (preservation of cognition and physical function after age 80).^13–17^ The effect of a protective allele may depend on the individual risk for developing dementia, due to advanced age or presence of AD-associated genetic variants. Ordered-subsets analysis (OSA) is an analytical tool that can identify increased evidence of linkage in the presence of trait heterogeneity by arranging families in the order of a continuous trait-related covariate.^18^ OSA may be implemented during NPL to identify subsets of families that are maximally informative for linkage based on higher or lower values of a covariate, such as a genetic risk score. This procedure has previously been used to detect AD loci, using age of onset as the ranking covariate.^19^ In this study, we examine Amish sibships enriched with aged, cognitively unimpaired (CU) individuals, to investigate shared genetic variants that promote cognitive preservation. We conducted a genome-wide linkage screen in a subset of families containing multiple CU members, followed by association tests of linked regions in a broader sample. Recognizing that cognitive preservation is a complex and heterogeneous trait, we calculated genetic risk scores (GRS) for each individual comprising previously identified genome-wide significant AD loci, weighted by effect sizes reported in Kunkle et al.^6^ These GRS were used to rank families in OSA analysis based on the average GRS of each CU individual, reflecting the relative degree of inherited AD risk. By integrating genetic risk of AD into a linkage analysis framework, we expected increased evidence of linkage and association in high-risk families enriched with CU individuals, possibly due to the presence of genetic variants buffering inherited AD risk.

## Materials and Methods

### Ascertainment

This study was conducted as a component of the Collaborative Amish Aging and Memory Project (CAAMP). CAAMP is a longitudinal study of cognitive function and age-related traits in the Midwestern Amish. Individuals participating in CAAMP belong to the communities in and around Adams, Elkhart, and LaGrange Counties in Indiana, as well as Holmes County in Ohio. Present-day Amish in these areas largely descend from European Amish emigrants that arrived between the 18^th^ and 19^th^ centuries.^12^

Since 2002, the CAAMP study has enrolled Amish participants throughout these communities. During this time, several ascertainment approaches have been followed. In earlier waves of recruitment, individuals were enrolled if they were aged ≥ 65 with at least one sibling showing signs of dementia, or ≥ 80 years and cognitively intact. DNA samples were collected for all available cognitively impaired relatives and all cognitively unimpaired relatives aged ≥ 80. Currently, community members aged ≥ 75 are considered for enrollment regardless of cognitive status.

### Genotyping and Imputation

Sample genotyping was performed on Illumina MEGAex+3k and GSA chip sets. Quality control (QC) for each set was carried out separately. Common QC steps included sample level filtering for genotype missingness (> 3%), and variant level selection for polymorphic SNPs. SNPs (MAF > 1%) strongly deviating from Hardy-Weinberg Equilibrium (HWE) expected frequencies (p < 1x10^-6^) were dropped. Mendelian errors were evaluated in closely related subjects; genotypes were excluded for all members of a nuclear family if they were flagged as inconsistent with Mendelian segregation.

The MEGAex+3k set contained 774 subjects after sample QC. A total of 2,038,867 SNPs and indels were included on the chip. All indels were dropped to improve imputation. SNPs were dropped if they were monomorphic, duplicated, or missing in > 5% of samples. In total, the working MEGAex+3k set contained 655,441 SNPs (chr 1-22, X).

The GSA set contained 1,322 subjects following sample QC. The initial array contained 712,189 SNPs and indels. All indels were dropped. SNPs were excluded if they were monomorphic, duplicated, or missing in > 10% of samples. The resulting set was checked for HWE deviations and Mendelian errors. The final, working GSA data contained 545,740 SNPs (chr 1-22, X).

The cleaned MEGAex+3k and GSA sets were individually imputed on the Michigan Server using the Haplotype Reference Consortium (HRC) reference panel (European population). Haplotypes were phased using the Eagle algorithm.^20^ After imputation, QC of each set was as follows: rare SNPs (MAF < 0.01) with an INFO score < 0.8 were dropped, common SNPs with an INFO score < 0.4 were dropped. The post-QC MEGAex+3k and GSA contained 8,781,203 and 9,201,299 SNPs, respectively. A merged set was prepared using SNPs present in both sets. The final, merged imputed dataset contained 2,096 samples, and continuously distributed genotype doses at 759,280 rare and 7,552,523 common SNPs.

### Sample Selection and Phenotype

Cognitive status for enrolled subjects was evaluated using the Modified Mini Mental Status Exam (3MS).^21^ The 3MS exam is an extension of the Mini Mental Status Exam, with improved sensitivity and specificity compared to the original test.^22^ The 3MS has a maximum score of 100 and provides a brief assessment of concentration, temporal orientation, spatial orientation, language, constructional praxis, abstract thinking, and list-generating fluency. In concordance with the Cache County Memory Study, we set an education-adjusted 3MS threshold of < 87 to designate likely cognitive impairment (CI).^23^ This cutoff has previously been described as optimally sensitive for detection of CI. At the time of this study, 1,855 genotyped subjects had at least one 3MS examination, with the total number of 3MS exams per individual ranging from zero to six (mean = 1.6). For those with multiple examinations, cognitive status was based on the most recent examination. A status of cognitively unimpaired (CU) was assigned to individuals > 74 years of age with an education-adjusted score > 86. Those with an education-adjusted 3MS score below 87 at any age were considered CI. Overall, 1,175 individuals met these criteria: 715 were CU and 460 were CI. The mean age of the CU and CI subjects was 81.7 (SD 3.9) and 81.8 (SD 7.4), respectively.

The mean 3MS score of the CU and CI was 93.4 (SD 3.8) and 74.2 (SD 14.1), respectively. Women were a majority of both the CU (59 %) and CI (58 %) groups. A summary of the sample characteristics is provided in Table 1. Sibships were analyzed for linkage if they contained ≥ 2 CU members (n = 143 sibships). The number of CU individuals per sibship ranged from 2-7 (mode = 2).

**Table 1.** Demographic features, cognitive status, and demographic factors for susceptibility to cognitive impairment in study sample.

| Variable | Overall N = 1,175<sup>†</sup> | Unimpaired N = 715<sup>†</sup> | Impaired N = 460<sup>†</sup> |
| --- | --- | --- | --- |
| **Age at Exam (Years)** |  |  |  |
| Mean (SD) | 81.8 (5.5) | 81.7 (3.9) | 81.8 (7.4) |
| Median | 81.0 | 81.0 | 82.0 |
| **Sex** |  |  |  |
| F | 685 (58%) | 419 (59%) | 266 (58%) |
| M | 490 (42%) | 296 (41%) | 194 (42%) |
| **Education-Adjusted 3MS Score** |  |  |  |
| Mean (SD) | 85.9 (13.2) | 93.4 (3.8) | 74.2 (14.1) |
| Median | 89.0 | 93.0 | 80.0 |
| **GRS** |  |  |  |
| Mean (SD) | 3.0 (0.7) | 2.9 (0.6) | 3.1 (0.7) |
| Median | 2.8 | 2.8 | 2.9 |
| **APOE Genotype** |  |  |  |
| 2\|2 | 1 (<0.1%) | 1 (0.1%) | 0 (0%) |
| 2\|3 | 104 (8.9%) | 67 (9.4%) | 37 (8.0%) |
| 2\|4 | 21 (1.8%) | 11 (1.5%) | 10 (2.2%) |
| 3\|3 | 769 (65%) | 488 (68%) | 281 (61%) |
| 3\|4 | 257 (22%) | 140 (20%) | 117 (25%) |
| 4\|4 | 23 (2.0%) | 8 (1.1%) | 15 (3.3%) |

<sup>†</sup>n (%)

### Linkage Analysis

Non-parametric linkage analysis (NPL) was formed using Merlin to calculate multipoint Kong and Cox LOD* scores using the Whittemore and Halpern S_pairs_ scoring function.^24,25^ Merlin implements the Kong and Cox linear model to look for small increases in allele sharing across families, as is often the case for complex traits. The linkage dataset was prepared in PLINK.^26^ The QC’d MEGAex+3k and GSA chip sets were merged by overlapping SNPs. A representative panel of SNPs in low linkage disequilibrium (LD) was obtained by filtering for MAF > 0.3, then pruning using a window size of 500 kb, step size of 5, and r^2^ threshold of 0.16. The final linkage dataset contained 2,584 autosomal SNPs spaced approximately 1 Mb apart.^27^

### Genetic Risk Scores

The software package PRSice2 was used to calculate a LOAD GRS for each sample.^28^ In Kunkle et al., 26 loci associated with LOAD were identified through a large-scale GWAS and meta-analysis in a European population. Given that the study exclusively utilized clinically diagnosed AD cases, included the effect of *APOE*, and was performed in a European-descended population similar to the Amish, the effect sizes reported by Kunkle et al. were applied to estimate the true effects in the Amish sample population. Odds-ratios (OR) < 1 were inverted so that each estimate was expressed with respect to the risk-increasing allele. Due to poor imputation quality, the HLA variant rs9271058 was substituted using a proxy variant in complete LD (rs9271098). In addition, the imputation did not result in variant calls passing quality thresholds for the rare *TREM2* LOAD risk allele in the overall sample, therefore this locus was analyzed as monomorphic for the common allele, which does not increase risk (OR = 1). GRS calculations utilized an additive model, taking the sum of effect-size weighted genotype doses for each LOAD risk locus. A summary of the GRS variants is provided in Supplementary Table 1.

### Ordered Subsets Analysis

Ordered-subsets analysis (OSA) was performed following linkage analysis using in-house scripts (OSA2, based on the original OSA program). OSA serves as an extension to linkage analysis designed to help identify linkage in a subset of families defined by a trait-related covariate. To summarize the procedure, multipoint LOD scores are calculated at each marker position for a given chromosome, starting with the first family. The maximum LOD* and corresponding position are recorded. The next family is added to the subset, and the maximum LOD* score and position are recorded again. This process continues until all families have been added, and the subset with the highest LOD* score is identified. The significance of the LOD* increase is assessed by permutation testing, using an adaptive procedure with up to 10,000 permutations to calculate an empirical p-value. OSA signals were considered significant if the maximized LOD* exceeded 3, with an empirical p-value of less than 0.05. Signals with a maximized LOD* between 2 and 3, and an empirical p-value of less than 0.05, were considered suggestive. For this study, the ranking covariate for OSA was defined as the mean GRS of the CU individuals in each sibship.

### Genetic Association Testing

The R/Bioconductor GENESIS package was used to perform single-variant association tests within a generalized linear mixed model framework.^29^ Variants located in regions exhibiting significant LOD* increases during OSA were evaluated. For each region of interest, we selected variants with a MAF > 0.01 positioned within a 1-LOD down support interval of the peak linkage signal. GENESIS functionalities include tools to infer and adjust for population structure and genetic kinship, fit a null model, and perform single or aggregate association tests. GENESIS uses PC-AiR to estimate population structure, which is adjusted for sample relatedness by first selecting an ancestry representative panel of unrelated individuals for principal component analysis (PCA), then computing principal components (PCs) for the remainder of the sample based on genetic similarity. Pairwise kinship estimates for the entire sample were generated using the PC-Relate method, which leverages ancestral principal components to provide robust kinship estimates in the presence of population structure.

All 1,175 individuals that met the phenotype criteria were analyzed. PC-AiR and PC-Relate objects were generated using an LD-pruned, genome-wide panel of genotyped SNPs. The null model was fit using CU status as the outcome and included fixed effects for age, sex, GRS, and PC’s 1 and 2, as well as a kinship matrix as a random effect. The full model was generated by the addition of a SNP dose term to the null model for each imputed genotype within the intervals of interest. The Bonferroni-adjusted significance threshold in each region was set using the SimpleM procedure, which estimates the effect number of independent test (M_eff_) in the presence of high LD.^30^

To investigate whether the effect of an allele on CU status varies depending on the degree of inherited risk, we performed an additional regional analysis that incorporated a GRS x SNP interaction term. For each region of interest, the GRS was dichotomized, where individuals with a GRS higher than the mean GRS of the final sibship in the corresponding OSA subset were classified as high GRS (value = 1), and those with a GRS below the mean were classified as low GRS (value = 0). Tests of interaction within the GENESIS package use a joint Wald test to assess whether adding both the SNP main effect and the interaction term improves the model fit. The null model was fit using the same fixed and random effect terms as in the previous analysis.

## Results

### Non-Parametric Linkage and Ordered Subsets Analysis

The results of the genome-wide, multipoint, affected-only NPL analysis of cognitively unaffected individuals (CU) are illustrated in Figure 1. Prior to OSA, the strongest evidence of linkage was observed at 37.4 Mb on chr 22 (LOD* = 1.72).

**Figure 1.**
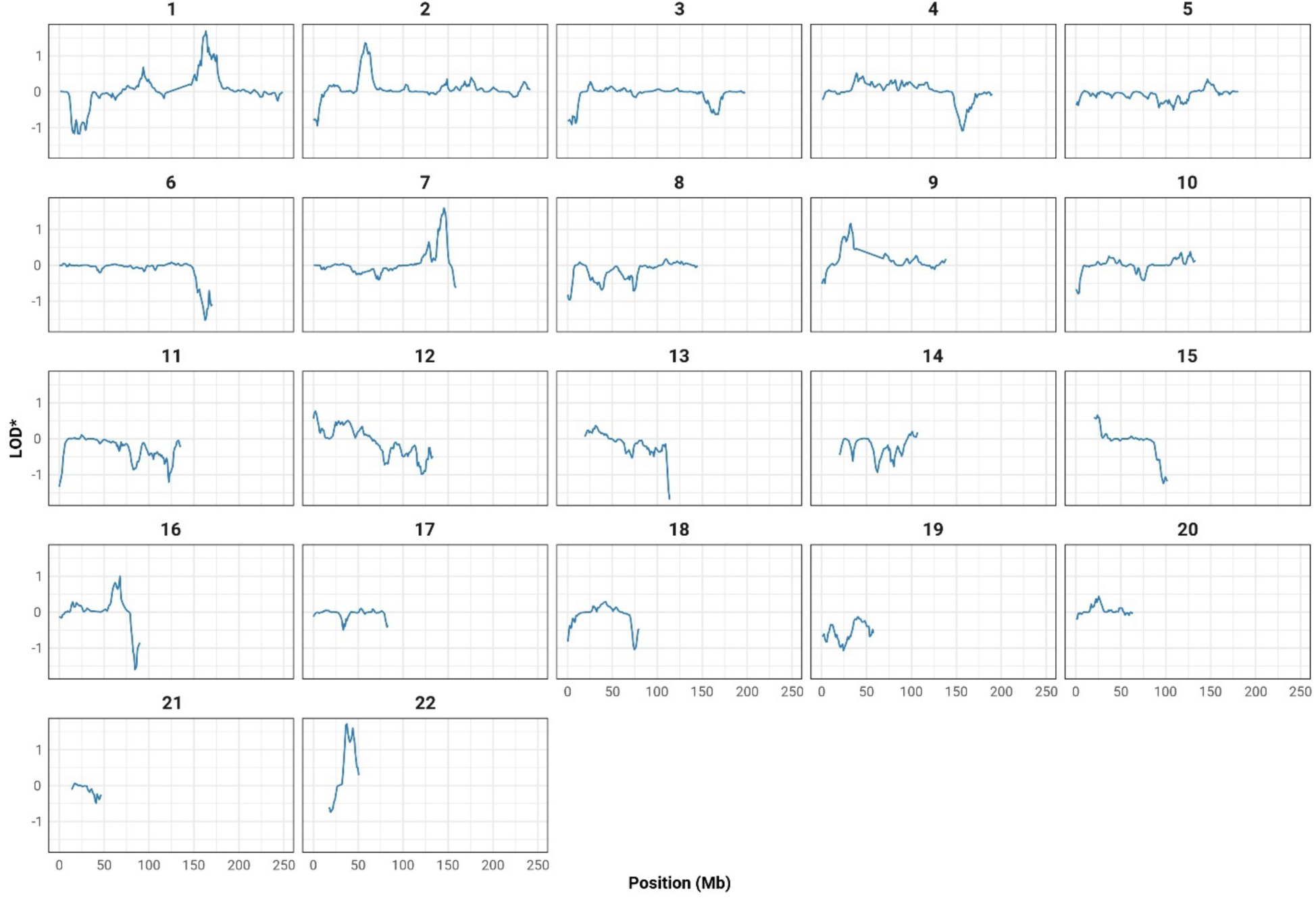
Non-Parametric Linkage Results Sibships with > 1 CU Member. **S_pair_** **N_Sibships_ = 143** **N_Unimpaired_ = 367**

The mean GRS for the overall sample (n = 1,175) was 3.0 (SD = 0.7), ranging from 1.7 to 5.5. During OSA, when ranking families from high to low mean GRS, we observed a significant LOD* increase (LOD* > 3, p_OSA_ < 0.05) on chromosome 2 (LOD* = 3.08, p_OSA_ = 0.043, 59.03 Mb, N = 79 families), as shown in Figure 2. Suggestive LOD* increases (2 < LOD* < 3, p_OSA_ < 0.05) were observed on chromosomes 3, 4, 6, and 18. The maximum OSA LOD* scores for these signals were: 2.48 (p_OSA_ = 0.044, 36.60 Mb, N = 40 families) on chromosome 3; 2.75 (p_OSA_ = 0.042, 64.40 Mb, N = 40 families) on chromosome 4; 2.38 (p_OSA_ = 0.035, 40.31 Mb, N = 26 families) on chromosome 6; and 2.8 (p_OSA_ = 0.0091, 47.16 Mb, N = 25 families) on chromosome 18.

**Figure 2.**
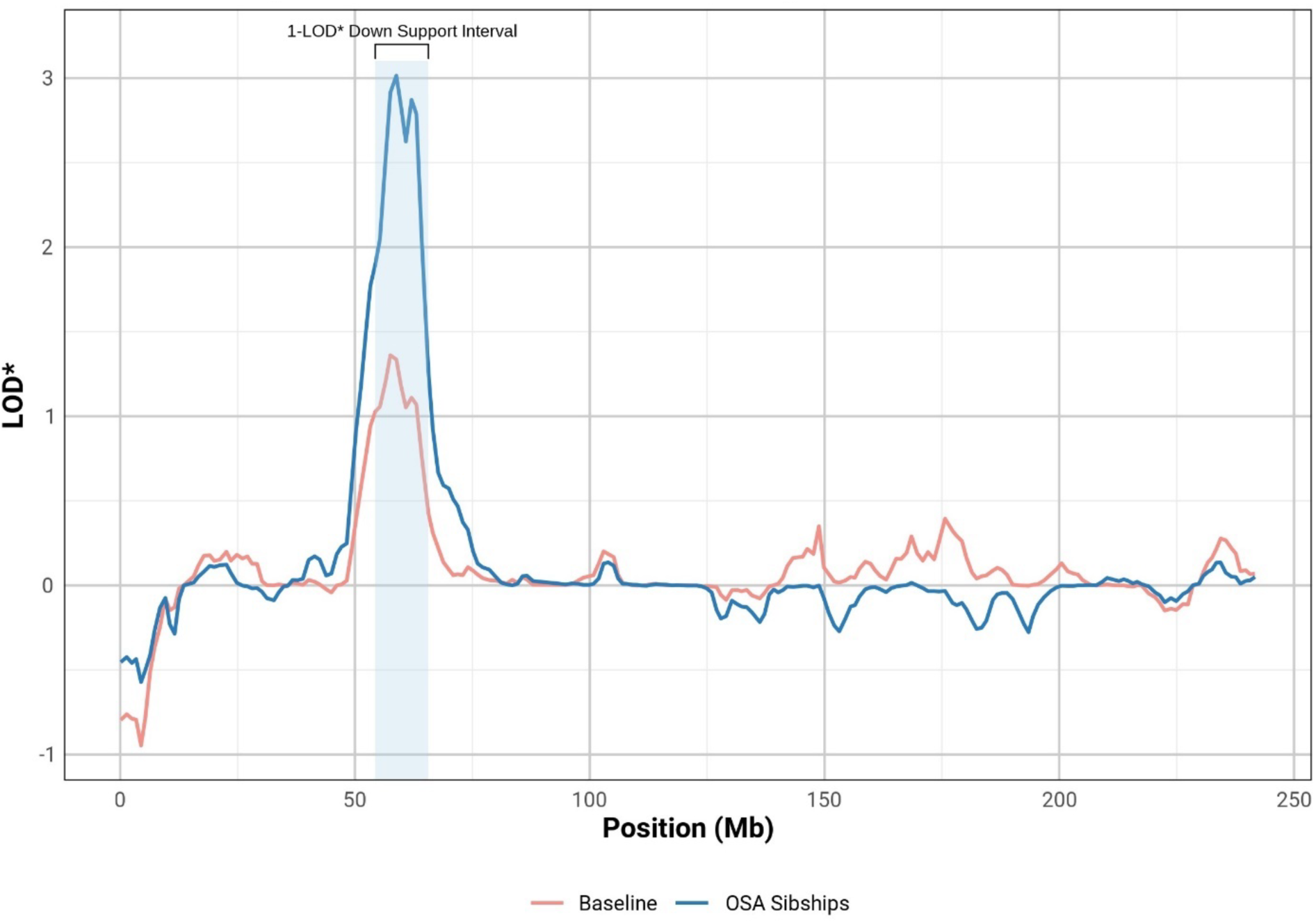
Comparison of Baseline and OSA Sibships on Chromosome 2.

Due to the disproportionately large effect of the *APOE* ε4 allele, the GRS tends to be higher, on average, in individuals who carry ε4. In OSA, this can result in an overrepresentation of ε4 carriers in the most at-risk sibships when using the GRS as the ranking covariate. When sibships are ordered by the mean GRS of cognitively intact sibship members, the first 20 families each contain at least one individual carrying an ε4 allele. To determine if ranking by mean GRS increased evidence of linkage in sibships without ε4, we repeated the OSA procedure in the subset of 95 sibships without any ε4 carriers. When ranking families by descending GRS, suggestive LOD* increases were observed on chromosomes 11 (LOD* = 2.46, p_OSA_ = 0.0036, 44.01 Mb, N = 12 families) and 14 (LOD* = 2.76, p_OSA_ = 0.0062, 69.29, N = 24 families). A maximum OSA LOD* score of 3.30 was observed on chromosome 7 (p_OSA_ = 0.20, 145.43 Mb), although this increase was not statistically significant. Additionally, we performed an OSA screen in the remaining subset of 48 sibships with at least one *APOE* ε4 carrier. These results mirrored the overall screen, with suggestive OSA LOD* increases observed on chromosomes 3 (LOD* = 2.50, p_OSA_ = 0.023, 36.60 Mb, N = 35 families), 4 (LOD* = 2.77, p_OSA_ = 0.020, 64.4 Mb, N = 16 families), 5 (LOD* = 2.21, p_OSA_ = 0.043, 66.51 MB, N = 12 families), 6 (LOD* = 2.38, p_OSA_ = 0.031, 40.31, N = 24 families), and 18 (LOD* = 2.57, p_OSA_ = 0.010, 47.16 Mb, N = 23 families). The OSA results are summarized in Table 2.

**Table 2.** Significant and suggestive OSA results for the overall, *APOE* ε4 positive, and *APOE* ε4 negative analyses.

| Position<sup>1</sup> | ID | P | Baseline LOD* | OSA LOD* | N Families | OSA Run | Function | Nearest Gene |
| --- | --- | --- | --- | --- | --- | --- | --- | --- |
| **Chr 2** |  |  |  |  |  |  |  |  |
| **59036916** | **rs6719884** | **0.043** | **1.36** | **3.08** | **79** | **all** | **ncRNA intronic** | ***LINC01122*** |
| **Chr 3** |  |  |  |  |  |  |  |  |
| 36596036 | rs1879545 | 0.023 | 0.65 | 2.50 | 35 | $\epsilon 4 +$ | intergenic | *STAC;DCLK3* |
| 36596036 | rs1879545 | 0.044 | 0.28 | 2.48 | 40 | all | intergenic | *STAC;DCLK3* |
| **Chr 4** |  |  |  |  |  |  |  |  |
| 64395080 | rs11737889 | 0.020 | 0.78 | 2.77 | 16 | $\epsilon 4 +$ | intergenic | *ADGRL3-AS1;TECRL* |
| 64395080 | rs11737889 | 0.042 | 0.52 | 2.75 | 16 | all | intergenic | *ADGRL3-AS1;TECRL* |
| **Chr 5** |  |  |  |  |  |  |  |  |
| 66513326 | rs1627990 | 0.043 | 0.53 | 2.21 | 12 | $\epsilon 4 +$ | intergenic | *CD180;LOC101928858* |
| **Chr 6** |  |  |  |  |  |  |  |  |
| 40312346 | rs13203076 | 0.031 | 0.69 | 2.37 | 24 | $\epsilon 4 +$ | ncRNA exonic | *LINC00951* |
| 40312346 | rs13203076 | 0.035 | 0.08 | 2.38 | 26 | all | ncRNA exonic | *LINC00951* |
| **Chr 11** |  |  |  |  |  |  |  |  |
| 44014531 | rs7103415 | 0.0036 | 0.00 | 2.46 | 12 | $\epsilon 4 -$ | intergenic | *C11orf96;ACCSL* |
| **Chr 12** |  |  |  |  |  |  |  |  |
| 2538549 | rs2239074 | 0.024 | 0.78 | 2.72 | 22 | $\epsilon 4 +$ | intronic | *CACNA1C* |
| **Chr 14** |  |  |  |  |  |  |  |  |
| 69751282 | rs7140574 | 0.0062 | 0.28 | 2.76 | 24 | $\epsilon 4 -$ | intronic | *GALNT16* |
| **Chr 18** |  |  |  |  |  |  |  |  |
| 47160814 | rs7228085 | 0.0091 | 0.29 | 2.83 | 25 | all | intergenic | *LIPG;ACAA2* |
| 47160814 | rs7228085 | 0.010 | 0.53 | 2.57 | 23 | $\epsilon 4 +$ | intergenic | *LIPG;ACAA2* |

<sup>1</sup>Build hg19.
Rows in bold indicate a significant LOD\* increase ( $p < 0.05$ ) and an OSA LOD\* $> 3$

### Regional Association Mapping

SNPs located within a 1-LOD down support interval of significant and suggestive OSA LOD* peaks observed during the three screens (combined, ε4 negative, ε4 positive) were tested for association with CU in the overall sample (N = 1,175). The genomic intervals, total loci, and the SimpleM-adjusted significance thresholds for each region are provided in Supplementary Table 2. Prior to fitting the SNP dosage term, the null model identified that GRS (p = 4.20 x 10^-5^) and PC1 (p = 1.96 x 10^-5^) were significantly associated with CU, while age (p = 0.63), sex (p = 0.71), and PC2 (p = 0.91) were non-significant.

#### Single Variant Analysis

A summary of the peak signals for each region tested is provided in Table 4. Association exceeding the adjusted significance thresholds was not observed. The strongest evidence of association overall was observed at the long intergenic non-coding RNA (lincRNA) SNP rs6740348 in *LINC02579* on chromosome 2 (p = 2.12 x 10^-5^, MAF = 0.15, OR = 0.57), located 5.81 Mb upstream of the OSA linkage peak. This signal was strongly correlated with 5 additional variants with p < 5 x 10^-5^ located within 5 kb. This locus is located approximately 20 kb upstream and downstream of genes *AFTPH* and *SERTAD2*, respectively. The strongest evidence of association relative to the regional significance threshold was observed at the intronic SNP rs33725 in *MAST4* on chromosome 5 (α_Adjusted_ = 1.05 x 10^-5^, p = 2.17 x 10^-5^, MAF = 0.22, OR = 0.62). This signal was also correlated with several other variants in the region with p < 5 x 10^-5^, including 14 intronic SNPs in *MAST4* and 12 intergenic variants near *CWC27* and *ADAMTS6*. The peak signal on chromosome 5 was located approximately 410 kb from the OSA LOD* peak identified during the ε4 carrier screen. The peak signal on chromosome 3 (rs75712982) was closest to its corresponding OSA peak, located approximately 250 kb away.

#### SNP x GRS_High_ Interaction Analysis

By incorporating the SNP x GRS interaction term, we observed significant evidence of an interaction effect at intergenic SNP rs11063479 on chromosome 12 (p_SNPxHIGH_GRS_ = 4.33 x 10^-6^, p_joint_ = 1.58 x 10^-5^). This SNP is located 4 kb upstream of *KCNA5*, which encodes the potassium voltage-gated, member 5 protein. During the initial regional analysis, rs11063479 was not associated with CU (p = 0.42). To further evaluate the effect of the variant across GRS strata, we stratified the sample into high (N = 273) and low (N = 902) risk using the corresponding OSA cutoff for chromosome 12 (GRS_high_ > 3.49) and inspected the result within each set. In the low GRS subset, rs11063479 was nominally associated with decreased odds of CU (p = 1.5 x 10^-3^, OR = 0.71). In contrast, the variant was more strongly associated with CU in the high GRS subset, with an opposite direction of effect (p = 2.2 x 10^-4^, OR = 2.07), indicating an increased odds of CU with increasing minor allele dosage. Each of the peak results from the initial regional analysis (Table 3) showed nominally significant joint p-values during the interaction analysis, however, the SNP x GRS interaction terms were non-significant. Therefore, the joint p-values for these tests were primarily driven by the relatively strong main effects. The results of the interaction analysis are summarized in Table 4.

**Table 3.** Summary of results for single-variant regional association tests within the 1-LOD intervals.

| Chr | ID | Position<sup>1</sup> | MAF | OR | P | Function | Nearest Gene | Peak LOD* Distance (Mb) |
| --- | --- | --- | --- | --- | --- | --- | --- | --- |
| 2 | rs6740348 | 64842018 | 0.15 | 0.57 | $2.12 \times 10^{-5}$ | ncRNA intronic | *LINC02579* | 5.81 |
| 3 | rs75712982 | 36349436 | 0.026 | 3.1 | $1.46 \times 10^{-4}$ | intergenic | *ARPP21*;<br>*STAC* | 0.25 |
| 4 | rs115327272 | 56093253 | 0.035 | 3.0 | $2.43 \times 10^{-4}$ | intergenic | *KDR*;<br>*SRD5A3* | 8.30 |
| 5 | rs33725 | 66098559 | 0.22 | 0.62 | $2.17 \times 10^{-5}$ | intronic | *MAST4* | 0.41 |
| 6 | rs10948196 | 44959384 | 0.37 | 0.68 | $7.67 \times 10^{-5}$ | intronic | *SUPT3H* | 4.65 |
| 11 | rs11038830 | 46262022 | 0.15 | 1.6 | $1.24 \times 10^{-4}$ | intergenic | *LOC101928894*;<br>*CREB3L1* | 2.25 |
| 12 | rs117138608 | 8599384 | 0.017 | 0.20 | $7.42 \times 10^{-4}$ | intergenic | *LINC00937*;<br>*CLEC6A* | 6.06 |
| 14 | rs35614357 | 71387980 | 0.13 | 1.7 | $7.17 \times 10^{-5}$ | intronic | *PCNX* | 1.64 |
| 18 | rs11663577 | 46397776 | 0.36 | 0.70 | $2.79 \times 10^{-4}$ | intergenic | *CTIF*;<br>*SMAD7* | 0.76 |

<sup>1</sup>Build hg19.

**Table 4.** Summary of results for SNP x GRS interaction tests within the 1-LOD intervals.

| Chr | ID | Position<sup>†</sup> | MAF | SNP x GRS<br>P | Joint P | Function | Nearest<br>Gene | Peak LOD* Distance<br>(Mb) |
| --- | --- | --- | --- | --- | --- | --- | --- | --- |
| 2 | rs11899888 | 56102744 | 0.17 | $1.06 \times 10^{-4}$ | $5.45 \times 10^{-4}$ | intronic | *EFEMP1* | 2.93 |
| 3 | rs73055959 | 36030320 | 0.023 | $1.46 \times 10^{-3}$ | $5.54 \times 10^{-3}$ | intergenic | *ARPP21*;<br>*STAC* | 0.57 |
| 4 | rs10008115 | 63330100 | 0.049 | $2.70 \times 10^{-4}$ | $4.60 \times 10^{-4}$ | intergenic | *ADGRL3-AS1*;<br>*TECRL* | 1.06 |
| 5 | rs79828202 | 68539253 | 0.055 | $4.10 \times 10^{-4}$ | $1.91 \times 10^{-3}$ | intronic | *CDK7* | 2.03 |
| 6 | rs1233421 | 29501514 | 0.47 | $2.16 \times 10^{-4}$ | $1.05 \times 10^{-3}$ | downstream | *LINC01015* | 10.81 |
| 11 | rs7943228 | 36229584 | 0.49 | $2.76 \times 10^{-4}$ | $8.43 \times 10^{-4}$ | intronic | *LDLRAD3* | 7.78 |
| 12 | rs11063479 | 5149103 | 0.29 | **$4.33 \times 10^{-6}$** | $1.58 \times 10^{-5}$ | intergenic | *KCNA1*;<br>*KCNA5* | 2.61 |
| 14 | rs35595126 | 65226044 | 0.13 | $2.18 \times 10^{-3}$ | $2.75 \times 10^{-3}$ | intronic | *SPTB* | 4.53 |
| 18 | rs28431791 | 31645148 | 0.30 | $8.36 \times 10^{-5}$ | $4.33 \times 10^{-4}$ | intronic | *NOL4* | 15.52 |

<sup>†</sup>Build hg19.
Cells highlighted in bold indicate statistically significant results relative to the SimpleM adjusted threshold

## Discussion

In this study, we identified significant evidence of linkage of CU to chromosome 2 using OSA, specifically at SNP rs6719884 (∼ 59.0 Mb) located in the lincRNA *LINC01122*. Additionally, a significant interaction effect was observed on chromosome 12 at SNP rs11063479 (∼ 5.1 Mb) near the *KCNA5* gene. These findings suggest that genetic factors within these regions may influence preserved cognitive function in individuals with a high genetic risk of Alzheimer’s disease (AD).

When considered as an outcome in and of itself, preserved cognitive function represents an etiologically complex trait that is driven by a combination of environmental and genetic factors. Unlike disease diagnoses, which typically adhere to established criteria for impairment, investigating preserved cognition—especially in relation to resilience to risk factors—requires context-specific approaches. Some studies in this field employ extreme sampling methods to explore the factors contributing to remarkably high cognitive functioning in later life. In this investigation, the primary neurocognitive assessment available for the majority of Amish participants was the 3MS. While the 3MS is highly sensitive for detecting cognitive impairment, it may lack the specificity to differentiate between varying levels of cognitive performance in individuals who are cognitively unimpaired. However, given that a high burden of inherited risk (e.g., high GRS) is associated with an increased likelihood of cognitive impairment, we hypothesized that high-risk sibships enriched with cognitively unimpaired individuals may be sharing genetic factors that buffer against this genetic risk. We tested this hypothesis using a modified linkage framework to identify excess allele sharing in families with a high AD risk, based on their mean GRS.

While we did not observe evidence of linkage in the overall dataset comprising 143 sibships, the OSA revealed significant evidence of linkage on chromosome 2 among CU individuals in sibships with a high GRS compared to the baseline. The peak LOD* score occurred at SNP rs671984 located in the lincRNA *LINC01122.* The biological function of *LINC01122* is poorly understood; however, variants in *LINC01122* are associated with a range of phenotypes, some of which include obsessive compulsive disorder, Tourette syndrome, insomnia, and episodic memory—a component of cognitive preservation.^31–33^ Additionally, GTEx expression data indicates that *LINC01122* is primarily expressed in the brain, particularly in the frontal cortex.^34^ Notably, while most of the OSA signals were driven by *APOE* ε4-positive and ε4-negative sibships, the observed linkage to chromosome 2 only occurred in the combined analysis. This suggests that a shared genetic factor within this region may influence preserved cognitive function in those with high genetic risk of AD, across *APOE* genotypes. It is worth noting that our research group recently reported evidence of linkage to cognitive preservation on chromosome 2 in an extended family with 24 unimpaired individuals.^35^ Comparing the two sets of results revealed that the linkage peaks are approximately 20 Mb apart, with only four individuals from the previous analysis included in the linked subset (containing 209 individuals) reported here. Additionally, conditioning the chromosome 2 regional association analysis on the peak linkage marker from the previous analysis (rs1402906) did not affect the results, suggesting that the signals are likely independent.

The remaining signals—which were suggestive based on a LOD* between 2 and 3 and an OSA p-value below 0.05—were primarily driven by the presence or absence of *APOE* ε4. Nevertheless, in both the ε4-positive and ε4-negative OSA analyses, suggestive evidence of linkage was observed in a limited subset of families characterized by elevated AD risk relative to the baseline. This underscores the potential utility of considering genetic risk strata within both ε4-positive and ε4-negative subgroups, as each may have its own protective loci.

Non-parametric linkage analysis is used to evaluate departures from expected levels of allele sharing within pedigrees. To determine if the regions of excess sharing observed among related CU individuals in the linkage-OSA screen contained alleles present at a higher frequency in CU versus CI, we performed case-control tests of association while controlling for GRS. While the strongest overall evidence of association was observed within *LINC02579* on chromosome 2, the effect was non-significant after correcting for multiple comparisons. The literature on *LINC02579* is scarce; however, it was recently reported as being significantly differently expressed in an *in vitro* study of dysregulated microRNAs and lincRNAs in the presence of amyloid beta.^36^ The strongest evidence of association, relative to the adjusted threshold, was observed in *MAST4* on chromosome 5, which encodes a member of the microtubule-associated serine/threonine protein kinases. A earlier GWAS of hippocampal volume identified novel variants in *MAST4* as being associated with decreased hippocampal volume, as well as an increased risk of AD based on LD score regression.^37^ This peak variant is within 410 kb of the corresponding OSA peak, and the estimated effect suggests a reduced odds of being cognitively unimpaired with increasing minor allele dosage.

We also performed tests of interaction to evaluate whether variant effects significantly differed between “high” and “low” GRS strata, defined by a threshold identified by OSA analysis on each chromosome. This stratification tests whether certain alleles exhibit different effects depending on the overall genetic risk profile of individuals. Such differences may occur if an allele provides a ‘buffering’ effect in the presence of high genetic risk, similar to the protective effect observed with the Klotho-VS variant in APOE ε4 carriers. The joint Wald test implemented in GENESIS evaluates the combined effects of the SNP and interaction term simultaneously. This test is particularly powerful for detecting interactions where the effect of a variant differs across levels of an effect modifier—in this case, the GRS strata. It is most sensitive to when the directions of effect are opposite across strata, which may cancel each other out when analyzed in the combined dataset. In our analysis, a significant interaction effect was observed at SNP rs11063479 near *KCNA5*, a potassium channel gene that has been previously associated with atrial fibrillation and cardiac arrest.^38^ The high-GRS cutoff applied to the analysis of the chromosome 12 region contained 265 out of the 301 total *APOE* ε4 carriers in the complete dataset, suggesting that the observed interaction may be influenced by *APOE* status. The estimated effect of the variant showed increased odds of being cognitively unimpaired in the high genetic risk strata (OR = 2.07, p = 2.2 x 10^-4^) and a decreased odds in the lower risk strata (OR = 0.71, 1.5 x 10^-3^). When stratified by *APOE* status, a similar but less significant effect was observed in both ε4 carriers (OR = 1.75, 1.5 x 10^-3^) and ε4 non-carriers (OR = 0.71, 2.2 x 10^-3^). This suggests a potential synergistic interaction between the variant and aggregate genetic risk, which includes *APOE* and additional risk loci. Atrial fibrillation is a common form of arrhythmia associated with an increased risk of stroke, reduced cerebral blood flow, and subsequent cognitive decline.^39^ Studies have also linked atrial fibrillation to a higher risk of cognitive decline, independent of stroke, suggesting direct effects on brain function.^40^ It is plausible that genetic variants, such as those near *KCNA5*, could the increase risk of cognitive impairment through intermediate cardiovascular phenotypes, such as atrial fibrillation. Given that the observed variant rs11063479 is intergenic but located approximately 4 kb from *KCNA5*, it may affect regulatory elements controlling gene expression. Follow-up studies are required to further explain the potential mechanisms underlying this association. After a thorough review of the literature, the loci highlighted here have not been previously reported in association with Alzheimer’s disease or cognitive preservation.

This study highlights the complexity of cognitive function in older age, showing it to be a heterogeneous trait shaped by various factors. Our objective was to identify a more homogeneous subgroup within the cognitively unimpaired population who are at high genetic risk of impairment. Resilience refers to individuals with better-than-expected cognitive performance relative to their level of brain pathology or risk of disease, potentially due to compensatory mechanisms that enable the brain to adapt to injury.^41^ This resilience is crucial for maintaining cognitive function despite genetic and environmental risk factors. Although we do not have direct measures of brain pathology, it is possible that individuals at the highest risk strata—defined by the presence of *APOE* ε4 and additional risk alleles of modest effect—might possess other variants that support resilient responses. Some of the loci highlighted in this study could represent such variants. Overall, this study contributes to the growing body of literature on preserved cognitive function at older ages by identifying genetic factors that may protect against cognitive decline.

## Data Availability

The data referenced in this manuscript is available upon request and with principal investigator approval by reaching out to the primary author.

