## Supplementary Material for "Linkage and association of preserved cognitive function in the Midwestern Amish at higher genetic risk of Alzheimer disease"

### Supplementary Table 1. Summary of variants and effect sizes for genome-wide significant Alzheimer's disease risk variants from Kunkle et al. (2019) utilized in GRS calculation

| Chromosome | ID | Position[1] | Gene | Non-Effect Allele | Effect Allele | OR[2] | Notes |
|---|---|---|---|---|---|---|---|
| 1 | rs4844610 | 207802552 | *CR1* | C | A | 1.17 | - |
| 2 | rs6733839 | 127892810 | *BIN1* | C | T | 1.20 | - |
| 2 | rs10933431 | 233981912 | *INPP5D* | G | C | 1.09 | - |
| 6 | rs9271058 | 32575406 | *HLA-DRB1* | T | A | 1.10 | Not imputed; substituted with LD proxy rs9271098 in GRS |
| 6 | rs114812713 | 41034000 | *OARD1* | G | C | 1.32 | - |
| 6 | rs75932628 | 41129252 | *TREM2* | C | T | 2.08 | Not imputed; no suitable LD proxy, omitted from GRS |
| 6 | rs9473117 | 47431284 | *CD2AP* | A | C | 1.09 | - |
| 7 | rs12539172 | 100091795 | *NYAP1g* | T | C | 1.09 | - |
| 7 | rs10808026 | 143099133 | *EPHA1* | A | C | 1.11 | - |
| 8 | rs73223431 | 27219987 | *PTK2B* | C | T | 1.10 | - |
| 8 | rs9331896 | 27467686 | *CLU* | C | T | 1.14 | - |
| 10 | rs7920721 | 11720308 | *ECHDC3* | A | G | 1.08 | - |
| 11 | rs3740688 | 47380340 | *SPI1h* | G | T | 1.09 | - |
| 11 | rs7933202 | 59936926 | *MS4A2* | C | A | 1.12 | - |
| 11 | rs3851179 | 85868640 | *PICALM* | T | C | 1.14 | - |
| 11 | rs11218343 | 121435587 | *SORL1* | C | T | 1.25 | - |
| 14 | rs17125924 | 53391680 | *FERMT2* | A | G | 1.14 | - |
| 14 | rs12881735 | 92932828 | *SLC24A4* | C | T | 1.09 | - |
| 15 | rs593742 | 59045774 | *ADAM10* | G | A | 1.08 | - |
| 16 | rs7185636 | 19808163 | *IQCK* | C | T | 1.09 | - |
| 16 | rs62039712 | 79355857 | *WWOX* | G | A | 1.16 | - |
| 17 | rs138190086 | 61538148 | *ACE* | G | A | 1.30 | - |
| 19 | rs3752246 | 1056492 | *ABCA7* | C | G | 1.15 | - |
| 19 | rs429358 | 45411941 | *APOE* | T | C | 3.32 | - |
| 20 | rs6024870 | 54997568 | *CASS4* | A | G | 1.14 | - |
| 21 | rs2830500 | 28156856 | *ADAMTS1* | A | C | 1.08 | - |

[1]Build hg19

[2]Odds ratio relative to the effect allele

Variants and effect sizes as reported by Kunkle et al. (2019)

**Supplementary Table 2. Overview of genomic intervals, total loci, and independent tests for 1-LOD regions assessed in regional association follow-up**

| Chr | Peak Linkage Position[1] | Start Position[1] | End Position[1] | Total Loci | Independent Tests[2] | Adjusted Alpha[3] |
|---|---|---|---|---|---|---|
| 2 | 59036916 | 54562012 | 65877962 | 28151 | 6423 | $7.78 \times 10^{-6}$ |
| 3 | 36596036 | 35536237 | 46781836 | 27025 | 5769 | $8.67 \times 10^{-6}$ |
| 4 | 66421666 | 55049751 | 70350006 | 47611 | 9167 | $5.45 \times 10^{-6}$ |
| 5 | 66513326 | 61978989 | 73146026 | 22997 | 4779 | $1.05 \times 10^{-5}$ |
| 6 | 40312346 | 24590547 | 45722988 | 78753 | 15882 | $3.15 \times 10^{-6}$ |
| 11 | 44014531 | 35165138 | 55681336 | 46231 | 8581 | $5.83 \times 10^{-6}$ |
| 12 | 2538549 | 213998 | 9876091 | 26675 | 7181 | $6.96 \times 10^{-6}$ |
| 14 | 69751282 | 64788644 | 74227246 | 24221 | 5094 | $9.82 \times 10^{-6}$ |
| 18 | 47160814 | 30166313 | 57197019 | 70145 | 15459 | $3.23 \times 10^{-6}$ |

[1]Build hg19.

[2]Independent Tests based on SimpleM calculation.

[3]Adjusted Alpha = 0.05 / Independent Tests
